# Initial SARS-CoV-2 viral load is associated with disease severity: a retrospective cohort study

**DOI:** 10.1101/2021.10.01.21264412

**Authors:** Dennis Souverein, Karlijn van Stralen, Steven van Lelyveld, Claudia van Gemeren, Milly Haverkort, Dominic Snijders, Robin Soetekouw, Erik Kapteijns, Evelien de Jong, Gonneke Hermanides, Sem Aronson, Alex Wagemakers, Sjoerd Euser

## Abstract

**Background:** We aimed to assess the association between initial SARS-CoV-2 viral load and the subsequent hospital and intensive care unit (ICU) admission and overall survival.

**Methods:** All persons with a positive SARS-CoV-2 RT-PCR result from a combined nasopharyngeal (NP) and oropharyngeal (OP) swab (first samples from unique persons only) that was collected between March 17, 2020, and March 31, 2021, in Public Health testing facilities in the region Kennemerland, province of North Holland, the Netherlands were included. Data on hospital (and ICU) admission were collected from the two large teaching hospitals in the region Kennemerland.

**Results:** In total, 20,207 SARS-CoV-2 positive persons were included in this study, of whom 310 (1.5%) were hospitalized in a regional hospital within 30 days of their positive SARS-CoV-2 RT-PCR test. When persons were categorized in three SARS-CoV-2 viral load groups, the high viral load group (Cp < 25) was associated with an increased risk of hospitalization as compared to the low viral load group (Cp > 30) (ORadjusted [95%CI]: 1.57 [1.11-2.26], p-value=0.012), adjusted for age and sex. The same association was seen for ICU admission (ORadjusted [95%CI]: 7.06 [2.15-43.57], p-value=0.007). For a subset of 243 of the 310 hospitalized patients, the association of initial SARS-CoV-2 Cp-value with in-hospital mortality was analyzed. The initial SARS-CoV-2 Cp-value of the 17 patients who deceased in the hospital was significantly lower (indicating a higher viral load) compared to the 226 survivors: median Cp-value [IQR]: 22.7 [3.4] vs. 25.0 [5.2], OR[95%CI]: 0.81 [0.68-0.94], p-value = 0.010.

**Conclusions:** Our data show that higher initial SARS-CoV-2 viral load is associated with an increased risk of hospital admission, ICU admission, and in-hospital mortality. We believe that our findings emphasize the added value of reporting SARS-CoV-2 viral load based on Cp-values to identify persons who are at the highest risk of adverse outcomes such as hospital or ICU admission and who therefore may benefit from more intensive monitoring.

## INTRODUCTION

During the COVID-19 pandemic, the primary method to diagnose SARS-CoV-2 was molecular testing of respiratory samples by reverse transcriptase-polymerase chain reaction (RT-PCR). Although SARS-CoV-2 RT-PCR results generally are reported in a qualitative manner (positive or negative), the quantitative test result (cycle threshold (Ct), or crossing point (Cp)) indicates the viral load. This viral load may be related to disease phenotype, morbidity, and mortality of COVID-19 patients.

To date, several studies have investigated the association between viral load with different clinical parameters and outcome measurements in COVID-19 patients. Although some of these studies found a clear relationship between higher SARS-CoV-2 viral load and hospital admission, duration of hospital stay, or mortality,^1-3^ others found no^4^ or even an inverse relationship between higher viral load and clinical symptoms, hospitalization, ICU admission, and overall mortality.^5^ Differences between the results of these studies may be explained by the setting of the studies; most were restricted to hospitalized patient populations and used the SARS-CoV-2 RT-PCR test result performed at admission to the hospital to study relations between viral load and clinical outcomes. Because the viral load is highest in the earliest (mostly pre-symptomatic) stages of the infection, followed by a gradual decline, the moment of testing in the infection phase probably will influence the viral load found in respiratory samples. Therefore, a SARS-CoV-2 RT-PCR test result performed at admission to the hospital may not provide a representative estimate of the SARS-CoV-2 viral load in the early stage of the infection, thereby explaining the inconsistent results found in these previous studies.

In the Netherlands, like in many European countries, individuals with minor symptoms can have themselves tested at Public Health Services. The majority of patients who present at these facilities are in the early stage of the infection (median 2 days since start onset symptoms).^6^ In this study, we investigated whether initial SARS-CoV-2 viral loads of tests performed at a Public Health testing facility were associated with admission to regional hospitals and intensive care units (ICUs), and overall survival.

## METHODS

### Setting, study design, and participants

We performed a retrospective cohort study on data from persons with a positive SARS-CoV-2 RT-PCR result from combined nasopharyngeal (NP) and oropharyngeal (OP) swabs (first samples from unique persons only) that were analyzed between March 17, 2020, and March 31, 2021, using the RT-PCR based on the presence of the E-gene^7^ in the Regional Public Health Laboratory Kennemerland, Haarlem, the Netherlands. Cp-values were calculated on Lightcycler 480 1.5.1 software (Roche diagnostics, Basel, Switzerland). Only swabs derived from a Public Health testing facility in the region Kennemerland were included, as Public Health testing facilities employed a uniform sampling (combined naso- and oropharynx) and inclusion policy.^6^

Data on hospital (and ICU) admission were collected from the two large teaching hospitals in the region Kennemerland. These two hospitals are the only public hospitals in the region and the primary referral centers for patients from this region. Only data on hospitalization (and/or ICU admission) within 30 days from the date of the first positive SARS-CoV-2 RT-PCR result were taken into account. Data on hospital mortality were collected from one large teaching hospital by assessing electronic patient files.

### Ethics Statement

The Medical Ethical Committee of the Amsterdam UMC approved this study on January 19^th^, 2021 (Study number:2021.0170). Prior to this, the Institutional Review Board of the Spaarne Gasthuis, Hoofddorp/Haarlem, the Netherlands, approved the study (Study number: 2021.0046). The data were anonymized after collection and analyzed under code.

### Statistical analysis

Descriptive statistics were used to present the data: continuous variables were presented as median (interquartile range (IQR)), categorical variables were presented as numbers and percentages (%). Comparisons of continuous variables were made using independent sample t-tests or Mann-Whitney U tests (when applicable), for categorical variables Chi-square tests were used. Logistic regression analyses were used to analyze the relation between SARS-CoV-2 viral load and hospitalization, allowing additional adjustment for potentially confounding variables. Statistical analyses were performed with R and RStudio (R version 4.0.3) using packages tidyverse and tidymodels. P-values <0.05 were considered significant.

## RESULTS

A total of 20,207 individuals tested positive for SARS-CoV-2 in an RT-PCR test performed at a Public Health testing facility. Of these, 10,280 (50.9 %) were female, median [IQR] age was 40 [29] years and their median [IQR] SARS-CoV-2 Cp-value was 26.3 [5.7].

### Association between Cp-value and hospital admission

Of the 20,207 SARS-CoV-2 positive persons, 310 were hospitalized within 30 days of their positive SARS-CoV-2 RT-PCR test. Hospitalized patients were older (median age[IQR]: 64.0 [21.0] vs. 40.0 [30.0] years, OR [95%CI]: 1.07 [1.06-1.08], p-value<0.001) and more often male (60.0% vs. 49.0%, OR [95%CI]: 1.56 [1.25-1.97], p-value<0.001) compared with non-hospitalized persons.

Cp-values at time of baseline measurement were considerably lower (indicating higher viral loads) in those patients who were eventually hospitalized (median Cp-value [IQR]: 25.0 [5.2] vs. 26.3 [5.6], OR[95%CI]: 0.93 [0.90-0.96], p-value<0.001) as compared with persons who were not hospitalized. After adjusting for age and sex, the relation between initial SARS-CoV-2 viral load and hospitalization did not markedly change (ORadjusted [95%CI]: 0.96 [0.93-0.99], p-value = 0.004).

When patients were categorized in three SARS-CoV-2 viral load groups, the high viral load group (Cp < 25) was associated with an increased risk of hospitalization compared to the low viral load group (Cp > 30) (OR[95%CI]: 2.04 [1.46-2.93], p-value<0.001). This association did not markedly change after adjusting for age and sex (ORadjusted [95%CI]: 1.57 [1.11-2.26], p-value=0.012) (Supplemental Table 1). Analyzing the association of initial SARS-CoV-2 viral load and hospitalization in different age groups (Supplemental Table 5) showed a consistent association (although numbers per age group were limited) in all age groups. Additional data on the association between initial SARS-CoV-2 viral load and hospitalization over time (March 2020-March 2021) are presented in Supplemental Table 6. These show that the association was seen in all months, with the clearest association from October 2020 onwards (when the majority of tests was performed).

### Association between Cp-value and hospital Intensive Care Unit admission

There were 60 patients who were admitted to the ICU within 30 days of their initial positive SARS-CoV-2 RT-PCR test. Patients admitted to the ICU had lower initial SARS-CoV-2 Cp-values (indicating higher viral loads) compared to patients who were not admitted to the ICU (median Cp-value [IQR]: 24.5 [4.2] vs. 26.3 [5.7], OR[95%CI]: 0.87 [0.81-0.93], p-value<0.001). In addition, patients admitted to the ICU were older (median age [IQR]: 64.5 [13.2] vs. 40.0 [29.0] years, OR[95%CI]: 1.07 [1.06-1.09], p-value<0.001), and more often male (65.0% vs. 49.1%, OR[95%CI]: 1.93 [1.15-3.33], p-value = 0.016).

Adjusting the analysis on the relation between initial SARS-CoV-2 viral load and ICU admission for age and sex did not markedly change these results (ORadjusted [95%CI]: 0.90 [0.84-0.96], p-value = 0.004). The high viral load group (Cp < 25) was associated with an increased risk of ICU admission compared to the low viral load group (Cp > 30) (ORcrude [95%CI]: 9.34 [2.85-57.55, p-value=0.002), also after adjustment for age and sex (ORadjusted [95%CI]: 7.06 [2.15-43.57], p-value=0.007) (Supplemental Table 2).

### Association between Cp-value and clinical parameters in hospitalized patients

For a subset of 243 of the 310 hospitalized patients, data were available on in-hospital mortality. Of these 243 hospitalized patients, the association of their initial SARS-CoV-2 Cp-values with in-hospital mortality was analyzed. In total, 17 (7.0%) of these hospitalized patients died during their admission. The initial SARS-CoV-2 Cp-value of these 17 deceased patients was significantly lower (indicating higher viral load) compared to the 226 survivors: median Cp-value [IQR]: 22.7 [3.4] vs. 25.0 [5.2], OR[95%CI]: 0.81 [0.68-0.94], p-value = 0.010. In addition, these 17 patients were older (median age [IQR]: 79.0 [9.0] vs. 63.0 [20.0] years, OR[95%CI]: 1.12 [1.07-1.20], p-value < 0.001), and had more often been admitted to the ICU (52.9% vs. 16.8%, OR[95%CI]: 5.57 [2.01-15.73], p-value <0.001) compared to the patients who survived. There were no significant differences in sex distribution (proportion of males: 70.6% vs. 60.6%, OR[95%CI]: 1.69 [0.39-9.08], p-value = 0.502), BMI (median BMI [IQR]: 29.4 [3.6] vs. 26.6 [6.7], OR[95%CI]: 1.07 [0.92-1.23], p-value = 0.345) (Supplemental Table 3).

Adjusting the analysis on the relation between initial SARS-CoV-2 viral load and in-hospital mortality for age, sex, ICU admission, and BMI did not markedly change these results (ORadjusted [95%CI]: 0.81 [0.63-0.99], p-value = 0.062).

When the 243 patients who were included in this study (and for whom data were available on mortality) were compared to all other COVID-19 hospitalized patients from the participating hospitals (n=1,209) during the study period, the study patients were significantly younger (median age [IQR]: 65.0 [21.0] vs. 68.0 [22.0], OR[95%CI]: 0.99 [0.98-0.99], p-value<0.001) and showed a lower in-hospital mortality rate (proportion in-hospital mortality: 7.0% vs. 12.4%, OR[95%CI]: 0.53 [0.30-0.87], p-value = 0.017) (Supplemental Table 4).

## DISCUSSION

In this study of 20,207 SARS-CoV-2 positive persons who were tested in Public Health testing facilities, we found that lower SARS-CoV-2 RT-PCR Cp-values (indicating higher viral load) were associated with admission to hospital wards (Cp < 25 group compared to the low viral load group (Cp > 30), (ORadjusted[95%CI]: 1.57 [1.11-2.26], p-value=0.012) as well as to the ICU (ORadjusted [95%CI]: 7.06 [2.15-43.57], p-value=0.007). In addition, higher initial SARS-CoV-2 viral load was associated with in-hospital mortality.

Although several studies have investigated the association between viral load with different clinical parameters and outcome measurements in COVID-19 patients, a consensus has not been reached.^8^ Findings in studies that show an association of higher SARS-CoV-2 viral load and hospital admission, disease severity, or mortality,^1-3^ are in contrast with studies that found no or an inverse association.^4,5^ However, it should be noted that few studies analyzed>500 persons, the majority of them included only hospitalized patients, and almost none of them took into account that the moment of testing in the infection phase probably will influence the viral load found in respiratory samples.^8^

Our study is unique, as we have a large study population (20,207 SARS-CoV-2 positive persons), who were tested with a homogeneous sampling method. By only including tests performed by the Public Health Service we tried to remove heterogeneity of inclusion criteria, although the testing policy changed slightly during the study period (Supplemental text). As additional analysis showed that the association of higher viral load with hospital admission was stable over time (Supplemental Table 6), we do not think that the changes in testing policy have substantially affected our results. Furthermore, we anticipated on one of the problems that occur when studying SARS-CoV-2 viral loads in respiratory samples: the lack of comparability of Ct- or Cp-values derived from different laboratories, as these are assay- and method-specific.^9^ All samples in our study were analyzed in the same laboratory, using the same method.

One other distinguishing feature of our study is our study population of persons identified by the Public Health Services, where individuals can have themselves tested with minor symptoms. As the majority of patients who present at these facilities are in their early stage of the infection (median 2 days since start onset symptoms),^6^ the initial SARS-CoV-2 viral load assessed in these patients is more likely to represent a value close to the individual peak viral load compared to samples taken at a later stage of the disease (which is more common in studies that included hospitalized patients only.^8^

As has been postulated based on other viral infections, higher SARS-CoV-2 viral load may lead to increased disease severity and could by itself be a negative prognostic value for the course of the disease. Higher viral loads after infection could be the result of high inoculum doses^10^ and indeed, a dose-dependent effect of SARS inocula on morbidity and mortality was found in BALB/c mice.^11^ One may suggest that patients with higher SARS-CoV-2 inocula have higher initial viral loads, increasing the risk of hospital and ICU admission. Alternatively, one could postulate that higher initial viral loads are not necessarily caused by higher inocula, but rather by a reduced capability to clear the initial viral infection, leading to higher loads in the early stage of infection. Further studies should focus on this distinction. Of note, our study included persons before large-scale vaccination programs were in place, thus eliminating vaccination status as a cause for the correlation between higher viral loads and poor outcomes.

Our study has a few notable limitations. Although this is one of the first studies to connect viral load with increased risk of mortality, the number of in-hospital deaths in our study population was low, and larger studies are needed to assess the role of viral load in the outpatient setting and after adjustments for potential confounding factors. The data on hospital (and ICU) admission of our study population were collected from the two large teaching hospitals our region that largely cover the adherence area of the Regional Public Health Laboratory Kennemerland (where the tests were performed). It can however not be excluded that (ICU)hospitalization data of some of the included patients were missed when they were admitted to other hospitals in adjacent regions. However, we do not think that this will have influenced our main results as the chance of admission to a hospital in another region is not likely to be related to the initial SARS-CoV-2 viral load of a particular patient and would only have resulted in nondifferential misclassification of our outcome measurement. And finally, including only patients who were able to have themselves tested at Public Health Service testing facilities may have resulted in a healthy selection of all SARS-CoV-2 positive patients, as patients were able to make an appointment and go to the public health care facility. Even though this generally took place after a mean of 2 days, patients who got very ill, or needed to be admitted to the hospital may not be included. The data presented in Supplemental Table 4 seem to confirm a healthy selection of the patients, as the hospitalized study patients were significantly younger and showed lower in-hospital mortality rates compared to hospitalized patients not included in our study. This should be taken into account when interpreting the findings presented in this study.

In conclusion, we believe that our findings emphasize the added value of reporting SARS-CoV-2 viral load based on Cp-values in identifying persons who are at highest risk of adverse outcomes such as hospital or ICU admission and who therefore may benefit from more intensive monitoring. However, the interpretation of a single Ct- or Cp-value in less standardized settings should still be performed cautiously as it may be affected by sample collection, the time between the start of symptoms and sampling, and the lack of comparability of Ct- or Cp-values derived from different laboratories, as these are assay- and method-specific.^9^

## Data Availability

The data that support the findings of this study are not openly available and are available from the corresponding author upon reasonable request.

## Contributors

SE, SA, BH, AW and DS participated in conceptualisation. SE, SA, KS, SL, CG, DS, RS, EK, EJ, GH, AW and DS contributed to data collection. SE, AW and DS wrote the original draft. SA, CG, KS, SL contributed in reviewing and editing the paper. SE, KS, SA, AW and DS performed data curation and SE and DS contributed to data analysis. All authors critically reviewed and approved the final version.

## Acknowledgements

We would like to thank all physicians, nurses, Public Health testing personnel, laboratory technicians and administration personnel who have worked hard to provide SARS-CoV-2 testing for a large number of individuals, making it possible to perform these analyses.

---

**Figure 1.**
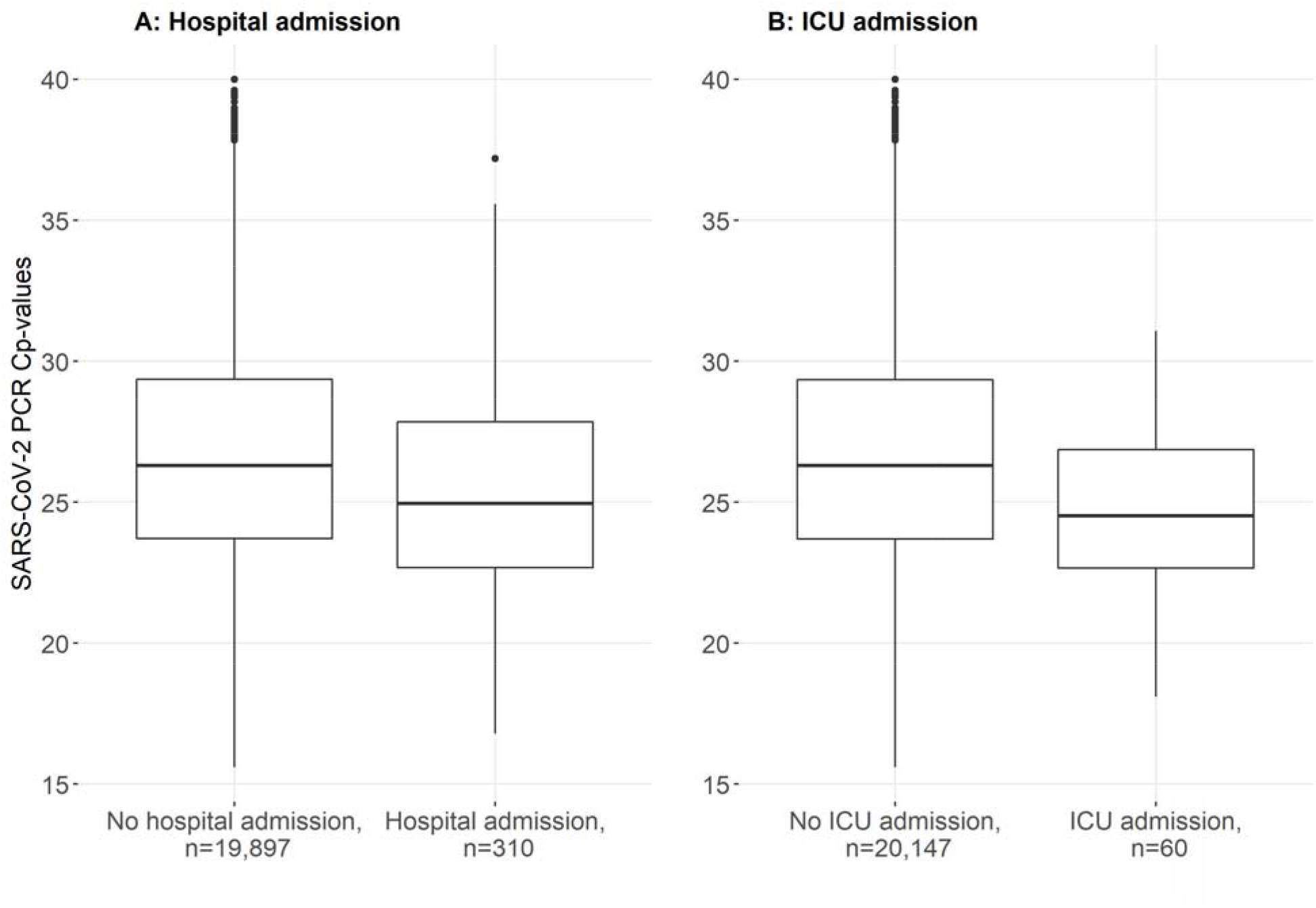
SARS-CoV-2 viral load distribution of 20,207 unique patients who were tested at a Public Health testing facility. Data on SARS-CoV-2 viral load distribution are presented for patients who were (n=310) or were not (n=19,897) admitted to hospital within 30 days after their initial SARS-CoV-2 RT-PCR test (panel A); and for patients who were (n=60) or were not (n=20,147) admitted to the ICU within 30 days after their initial SARS-CoV-2 RT-PCR test (panel B). Data are presented as Box-and-whisker plots with the central box covering the interquartile range with the median Cp-value indicated by the line within the box. The whiskers extend to the minimum and maximum values within 1.5 interquartile ranges of the quartiles, more extreme values are plotted individually.

## Supplementary material

**Supplemental table 1.** Association between baseline characteristics and hospital admission.

|  |  |  |  |  |  |  |  |
| --- | --- | --- | --- | --- | --- | --- | --- |
| **Baseline characteristics** | **Total population**<br>**(n=20.207)** | **Admission**<br>**(n=310)** | **No admission**<br>**(n=19.897)** | **OR univariable**<br>**[95%CI]** | **p-value** | **OR multivariable**<br>**[95%CI]** | **p-value** |
| Age, median [IQR] | 40.0 [29.0] | 64.0 [21.0] | 40.0 [30.0] | 1.07 [1.06-1.08] | <0.001 | 1.07 [1.06-1.08] | <0.001 |
| Male, No (%) | 9,927 (49.1) | 186 [60] | 9,741 [49.0] | 1.56 [1.25-1.97] | <0.001 | 1.50 [1.19-1.90] | <0.001 |
| Cp-value group |  |  |  |  |  |  |  |
| Cp-value >30 | 4,050 (20.0) | 41 [13.2] | 4,009 [20.1] | Ref | Ref | Ref | Ref |
| Cp-value 25-30 | 8,539 (42.3) | 113 [36.5] | 8,426 [42.3] | 1.31 [0.92-1.90] | 0.139 | 1.16 [0.81-1.69] | 0.417 |
| Cp-value <25 | 7,618 (37.7) | 156 [50.3] | 7,462 [37.5] | 2.04 [1.46-2.93] | <0.001 | 1.57 [1.11-2.26] | 0.012 |
| Data are presented as median[IQR] or No (%)percentages. IQR=interquartile range. Cp-value=crossing point value SD=standard deviation; Odds ratios (OR) and related p-values indicate the associations between baseline characteristics and hospital admission. |  |  |  |  |  |  |  |

**Supplemental table 2.** Association between baseline characteristics and ICU admission.

|  |  |  |  |  |  |  |  |
| --- | --- | --- | --- | --- | --- | --- | --- |
| **Baseline characteristics** | **Total population**<br>**(n=20.207)** | **ICU admission**<br>**(n=60)** | **No ICU admission**<br>**(n=20.147)** | **OR univariable**<br>**[95%CI]** | **p-value** | **OR multivariable**<br>**[95%CI]** | **p-value** |
| Age, median [IQR] | 40.0 [29.0] | 64.5 [13.2] | 40.0 [29.0] | 1.07 [1.06-1.09] | <0.001 | 1.07 [1.06-1.09] | <0.001 |
| Male, No (%) | 9,927 (49.1) | 39 [65.0] | 9,888 [49.1] | 1.93 [1.15-3.33] | 0.016 | 1.84 [1.09-3.20] | 0.025 |
| Cp-value group |  |  |  |  |  |  |  |
| Cp-value >30 | 4,050 (20.0) | 2 [3.3] | 4,048 [20.1] | Ref | Ref | Ref | Ref |
| Cp-value 25-30 | 8,539 (42.3) | 23 [38.3] | 8,516 [42.3] | 5.47 [1.62-34.08] | 0.021 | 4.86 [1.43-30.33] | 0.032 |
| Cp-value <25 | 7,618 (37.7) | 35 [58.3] | 7,583 [37.6] | 9.34 [2.85-57.55] | 0.002 | 7.06 [2.15-43.57] | 0.007 |
| Data are presented as median[IQR] or No (%)percentages. IQR=interquartile range. Cp-value=crossing point value SD=standard deviation; Odds ratios (OR) and related p-values indicate the associations between baseline characteristics and ICU admission. |  |  |  |  |  |  |  |

**Supplemental table 3.** Association between baseline characteristics and in-hospital mortality within a subset of admitted patients (n=243).

| Baseline characteristics | Total population<br>(n=243) | Died<br>(n=17) | Alive<br>(n=226) | OR univariable<br>[95%CI] | p-value | OR multivariable<br>[95%CI] | p-value |
| --- | --- | --- | --- | --- | --- | --- | --- |
| Age, median [IQR] | 65.0 [20.0] | 79.0 (9.0) | 63.0 (20.0) | 1.12 [1.07-1.20] | <0.001 | 1.21 [1.10-1.37] | <0.001 |
| Male, No (%) | 149 (61.3) | 12 (70.6) | 137 (60.6) | 1.56 [0.56-5.04] | 0.419 | 1.69 [0.39-9.08] | 0.502 |
| Cp-value, median [IQR] | 24.9 [5.2] | 22.7 [3.4] | 25.0 [5.2] | 0.81 [0.68-0.94] | 0.010 | 0.81 [0.63-0.99] | 0.062 |
| ICU admission, No (%) | 47 (19.3) | 9 (52.9) | 38 (16.8) | 5.57 [2.01-15.73] | <0.001 | 25.51 [5.22-184.76] | <0.001 |
| BMI, median [IQR] | 27.0 (6.6) | 29.4 [3.6] | 26.6 [6.7] | 1.04 [0.94-1.14] | 0.381 | 1.07 [0.92-1.23] | 0.345 |
| Data are presented as median[IQR] or No (%)percentages. IQR=interquartile range. Cp-value=crossing point value. Odds ratios (OR) and related p-values indicate the associations between baseline characteristics and in-hospital mortality. |  |  |  |  |  |  |  |

**Supplemental table 4.**
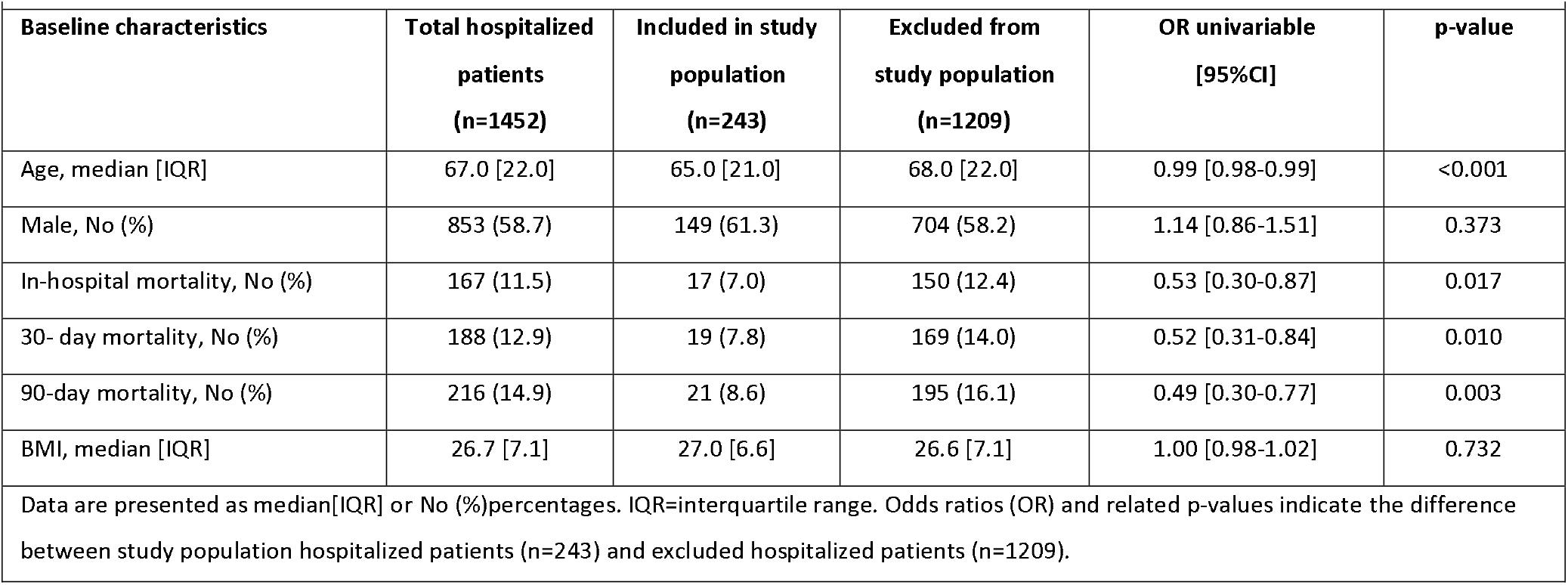
Differences between study population hospitalized COVID-19 patients (n=243) and excluded hospitalized COVID-19 patients (n=1,209).

**Supplemental table 5.** Cp-value characteristics for different age groups and sampling material within admitted (n=310) and not admitted patients (n=19897).

| Age groups | Admission | No admission | OR [95%CI] | p-value |
| --- | --- | --- | --- | --- |
| < 12 year | 1, 27.4 (0) | 692, 29 (5.1) | 0.88 [0.47-1.54] | 0.662 |
| 12-17 year | 3, 24.1 (5.8) | 1694, 27.2 (5.7) | 0.94 [0.69-1.25] | 0.697 |
| 18-29 year | 6, 23.1 (2.9) | 4190, 26.5 (5.8) | 0.78 [0.60-0.99] | 0.058 |
| 30-39 year | 22, 25.2 (3.3) | 3184, 26.5 (5.4) | 0.92 [0.81-1.02] | 0.134 |
| 40-49 year | 27, 25.4 (6.1) | 3420, 26.0 (5.5) | 1.07 [0.97-1.17] | 0.165 |
| 50-59 year | 64, 25.7 (5.2) | 3738, 25.8 (5.7) | 0.99 [0.93-1.06] | 0.849 |
| 60-69 year | 71, 26.0 (5.4) | 1809, 26.0 (5.5) | 0.97 [0.91-1.03] | 0.358 |
| 70-79 year | 79, 24.3 (4.4) | 840, 25.9 (5.9) | 0.91 [0.85-0.96] | 0.002 |
| > 79 year | 37, 24.3 (5.1) | 330, 25.5 (5.8) | 0.94 [0.85-1.02] | 0.155 |
| **Total** | **310, 25 (5.2)** | **19,897, 26.3 (5.6)** | **0.93 [0.90-0.96]** | **<0.001** |

Data are presented as No., median Cp-value (IQR). IQR=interquartile range. OR=Odds ratio. CI=confidence interval

**Supplemental table 6.** Cp-value characteristics for different moths during the study period within admitted (n=310) and not admitted patients (n=19897).

| Months | Admission | No admission | OR [95%CI] | p-value |
| --- | --- | --- | --- | --- |
| 2020.03 | 3, 20.8 (7.1) | 13, 24.9 (7) | 0.96 [0.71-1.19] | 0.704 |
| 2020.04 | 2, 32.1 (2.4) | 50, 29.1 (6.6) | 1.11 [0.81-1.55] | 0.510 |
| 2020.05 | 2, 30 (5.3) | 31, 27.7 (7.8) | 1.01 [0.79-1.54] | 0.588 |
| 2020.06 | - | 58, 26.8 (9) | - | - |
| 2020.07 | 2, 30.1 (2.3) | 142, 25.1 (6) | 1.28 [0.92-1.92] | 0.163 |
| 2020.08 | 3, 21.9 (7.7) | 527, 25.5 (6.9) | 1.03 [0.80-1.29] | 0.793 |
| 2020.09 | 17, 24.8 (7.2) | 2569, 25.9 (5.8) | 0.93 [0.82-1.05] | 0.272 |
| 2020.10 | 57, 23.1 (5.5) | 4964, 25.1 (5.5) | 0.87 [0.80-0.94] | <0.001 |
| 2020.11 | 41, 24.7 (4.2) | 2959, 26.1 (5.7) | 0.95 [0.87-1.02] | 0.184 |
| 2020.12 | 64, 24.9 (4.6) | 3837, 26.6 (5.6) | 0.87 [0.81-0.93] | <0.001 |
| 2021.01 | 34, 25.4 (4.4) | 1410, 27.3 (4.6) | 0.89 [0.80-0.98] | 0.022 |
| 2021.02 | 35, 26.2 (4.6) | 1539, 27.8 (4.6) | 0.89 [0.80-0.98] | 0.019 |
| 2021.03 | 50, 26.2 (4.8) | 1798, 27.8 (5.1) | 0.91 [0.80-0.98] | 0.017 |
| **Total** | **310, 25 (5.2)** | **19,897, 26.3 (5.6)** | **0.93 [0.90-0.96]** | **<0.001** |

Data are presented as No., median Cp-value (IQR). IQR=interquartile range. OR=Odds ratio. CI=confidence interval

## Supplemental text

### Nationwide SARS-CoV-2 PCR testing policy by Public Health (PH) authorities in The Netherlands

The most relevant changes in national testing policy in 2020 are summarized below. Testing by Public Health Services is free of charge.

### 27-01-2020

A coronatest can be performed when fulfilling the case definition:

Fever (≥ 38°C) plus 2 of the following symptoms: cough, runny nose, throat ache, pulmonary infiltrate) AND symptoms arose within 14 days of return from Wuhan or another area of local transmision or symptoms arose within 14 days of contact with a confirmesd COVID-19 patient.

From February 1^st^ ‘pulmonary infiltrate’ was substituted by ‘dyspnoea’.

### 07-02-2020

Case definition changed into:

Fever (≥ 38°C) plus 1 of the following symptoms: cough, dyspnea

AND symptoms arose within 14 days of return from China mainland (excluding Hong Kong, Macau and Taiwan) or symptoms arose within 14 days of contact with a confirmesd COVID-19 patient.

### 24-02-2020

Case definition changed into:

Fever (≥38°C) or feeling feverish in the elderly plus 1 of the following symptoms: cough, dyspnea (or any serious respiratory infection in the immunocompromised)

AND symptoms arose within 14 days of return from a country/region with widespread transmission or symptoms arose within 14 days of contact with a confirmesd COVID-19 patient.

### 06-03-2020

When family members of an index patient develop fever and/or respiratory symptoms they do not have to be tested, they will join in isolation. Only contacts with increased risk of severe disease (such as the elderly and immunocompromised) need to be tested.

### 12-03-2020

Case definition changed: The need for an epidemiological link to an area with widespread transmission or with a confirmed patient has been removed.

### 13-03-2020

Individuals fulfilling the case definition must stay at home until free of symptoms for 24 hours, they do not have te be tested. Outside hospitals, testing by PH is recommended only in persons with increased risk of more severe disease, who are suffering from fever AND respiratory symptoms (coughing or dyspnoea) and if this is of importance for their treatment. Increased risk is defined by age ≥ 70 years old or suffering from co-morbidities (judged by indication for yearly influenza vaccination).

### 19-03-2020

In (home) care institutions: after 1-2 proven cases of COVID-19 it is assumed there is an outbreak situation and additional patients are not tested.

### 06-04-2020

Health care workers who provide direct patient care, with symptoms for ≥ 24 hours consistent with COVID-19 (coughing and/or fever and/or rhinitis), can now be tested.

### 06-05-2020

The need to have symptoms ≥ 24 hours to be tested is removed. Paramedicals are now added to those eligible for testing. Children up to 12 years can be tested (upon approval of parents) when at least 3 children in their class/group are experiencing symptoms compatible with COVID-19.

### 01-06-2020

Everyone with symptoms of COVID-19 can be tested.

COVID-19 compatible symptoms are defined as a cold (such as runny nose, sneezing, throat ache) AND/OR coughing AND/OR sudden loss of taste or smell AND/OR dyspneoa AND/OR fever (>38°C). Contacts of index patients can be tested also in case of other COVID-19 symptoms such as headache.

### 13-08-2020 until 13-09-2020

Asymptomatic travellers from “code orange” countries / zones arriving at Amsterdam Airport Schiphol can temporarily be tested. The arrangement ends due to scarcity of testing material for which a national guideline in prioritizing in testing is necessary.

### 19-09-2020

Symptomatic children below the age of 13 are only tested when they are severely ill, are contact of an index patient or part of an outbreak investigation. This has several reasons, such as the low rates of positivity, lesser disease severity and the scarcity of testing material.

### 22-9-2020

If necessary children under the age of 6 and handicapped or psychogeriatric adults can be tested using saliva.

### 15-10-2020

Introduction of antigentests. Not used for health care personnel.

### 17-11-2020

Symptomatic children under 6 years are not by default tested. Children 6-12 years with only rhinitis can be tested, but it is not urgently adviced. Testing is urgently adviced when they are part of contact tracing, when they are severely ill (fever, dyspnoea or otherwise) or when testing is advised due to an outbreak investigation.

### 01-12-2020

Asymptomatic persons that are contacts of index patients can be tested at day 5 after the last contact.

### 03-02-2021

Asymptomatic persons that are a household or other close contact of index patients are urgently advised to get tested as soon as possible after contact, prior to the ‘day 5 after the last contact’ test.

### 19-2-2021

An antigentest can also be used to test contacts on or after day 5 of last contact.

